# Comorbidity Burden Predicts Patient-Important Adverse Outcomes and Informs Care Planning: A Retrospective Study from an Australian Metropolitan Hospital

**DOI:** 10.1101/2025.09.16.25335824

**Authors:** Hooi Hooi Koay, Mainak Majumdar, Umesh Kadam, Winnie Theresa, Sadeia Shah

## Abstract

**Background:** Mortality is a common clinical outcome indicator but fails to capture patient-important outcomes like functional independence, cognition, and quality of life.

**Objective:** To assess performance of three validated indices of comorbidity burden-Charlson Comorbidity Index (CCI), AHRQ Elixhauser Index (AHRQ-EI), Van Walraven modification (VW-EI)- for predicting patient-important adverse outcomes and healthcare resource utilisation.

**Methods:** Retrospective audit of all acute adult admissions to a metropolitan Australian teaching hospital submitted to the Victorian Admitted Episodes Dataset (VAED). Patient-important adverse outcomes were defined as a composite of in-hospital death, discharge to new residential care, or discharge against medical advice (DAMA). Data on ICU admission and hospital length of stay (LOS) was also collected. Receiver operating characteristic (ROC) curves were drawn to evaluate predictive accuracy of each score for composite adverse outcome.

**Results:** After excluding external transfers, 21,935 unique adult patients accounted for 29,055 separations (mean age 45.3 years; 79.6% female) were included. There were 356 in-hospital deaths (1.6%), with rates increasing by age and differing by gender.

Patients with prolonged hospitalisation (≥10 days, 90th percentile) had higher comorbidity scores than those with shorter stays (AHRQ-EI median 3 vs 0; VW-EI 5 vs 0; age-adjusted CCI 4 vs 0; all *p*<0.0001). Similarly, patients with repeated admissions (≥3 per year) had greater comorbidity burden compared with those with ≤2 admissions (AHRQ-EI median 0 vs 0; VW-EI 0 vs 0; age-adjusted CCI 2 vs 0; all *p*<0.0001). Patients admitted to ICU (n=491; 2.2%) showed the same pattern, with substantially greater comorbidity scores than non-ICU patients (AHRQ-EI median 8 vs 0; VW-EI 5 vs 0; age-adjusted CCI 3 vs 0; all *p*<0.0001). Despite representing only 2.2% of admissions, ICU patients accounted for 5.2% of hospital bed-days and had longer stays (median 5 vs 1 day; *p*<0.0001).

Among comorbidity indices, an age-adjusted CCI >4 had the strongest predictive performance for adverse outcomes (AUC 0.83; recall 0.70), followed by VW-EI >5 (AUC 0.77) and AHRQ-EI >5 (AUC 0.75). Despite elevated risk, 49% of patients aged ≥65 years and 36.5% of those with high comorbidity burden lacked documented goals-of-care (GOC) discussions.

**Conclusion:** Comorbidity burden is a strong predictor of patient-important adverse outcomes and increased healthcare resource use. Routine integration into workflows could trigger earlier discussions to better align care with patient values and inform treatment limitations where appropriate.

**KEY MESSAGES:** What is already known on this topic:

While mortality is a commonly used clinical outcome, it does not fully reflect outcomes that matter most to patients, such as independence and quality of life. Comorbidity indices have been used to predict mortality in healthcare but their application in guiding patient-centered care planning is less established.

What this study adds:

This retrospective audit shows that comorbidity burden, especially the age-adjusted Charlson Comorbidity Index, is a strong predictor of patient-important adverse outcomes and increased healthcare resource use.

How this study might affect research, practice or policy:

Routine use of comorbidity indices may help inform anticipatory care planning and prompt early discussions on goals-of-care discussions to improve alignment of care with patient values.

## INTRODUCTION

Prevention of death is the ultimate goal for many therapeutic interventions [1] and mortality is considered the “hardest”, most objective clinical outcome measure as it is unbiased and difficult to manipulate. Where survival is not the only concern [2], mortality may not fully capture outcomes that matter most to patients. For instance, long-term studies of Intensive Care Unit (ICU) survivors have demonstrated substantial disability and diminished health-related quality of life [3,4]. Patient-important outcomes, including maintenance of function, avoidance of premature institutionalisation, and quality of life, often outweigh survival alone in decision making [5,6]. Anticipating these outcomes is essential to guide shared decision making and aligning medical care with patient values for delivery of patient-centric care.

The Charlson Comorbidity Index (CCI) [7] was originally developed to predict 1-year mortality in longitudinal studies and has since been validated for longer-term outcomes including 10-year mortality. In contrast, the Elixhauser Comorbidity Index was initially introduced by Elixhauser et al. [8] to predict in-hospital outcomes using administrative data. This was subsequently modified by van Walraven et al. [9] into a weighted point system to enhance prediction of in-hospital mortality, and further adapted by the Agency for Healthcare Research and Quality (AHRQ) into the ICD-10-compatible AHRQ Elixhauser Index (AHRQ-EI) [10] for broader hospital-level use.

While these indices were developed primarily for mortality prediction, they have been increasingly applied to estimate additional hospital outcomes such as length of stay (LOS), hospital charges, and risk stratification [11–20]. Validated comorbidity indices like the CCI, AHRQ-EI, and VW-EI provide structured approaches to quantifying comorbidity burden and are widely used in risk adjustment and hospital performance evaluation. However, their utility in predicting broader patient-important outcomes and supporting anticipatory care planning in acute care settings remains less well established. We therefore aimed to evaluate the predictive performance of these indices for clinically meaningful outcomes using administrative data from the Victorian Admitted Episodes Dataset (VAED) for all acute adult admissions to a single Australian metropolitan teaching hospital in 2022.

## METHODS

### Selection and Description of Participants

A retrospective audit was conducted on all adult separations at a metropolitan public teaching hospital in Victoria, Australia, between January 1 and December 31, 2022. Hospital administrative data submitted to the VAED were used as the primary data source. The study population included all adult patients (aged ≥18 years) admitted to acute care services during the study period. To ensure accuracy in outcome assessment, patients transferred to other hospitals were excluded from the analysis. This decision was made as downstream hospital stay and discharge outcomes for these patients could not be reliably captured within the local dataset. ICU admissions were identified by cross-referencing internal ICU databases with scanned medical records. Demographic data, including age and sex, were collected for all included patients.

### Data Collection and Measurements

#### Comorbidity indices

Demographic and ICD-10-AM coded diagnosis-related group (DRG) data were entered into validated Excel-based comorbidity scoring tools [21] to compute three indices:

- The Charlson Comorbidity Index (CCI)
- The AHRQ Elixhauser Index (AHRQ-EI)
- The Van Walraven modification (VW-EI)

An age-adjusted version of the CCI was applied, incorporating patient age into the comorbidity score to enhance mortality risk prediction.

### Outcome measures

#### Primary outcome

The primary outcome was a composite measure of patient-important adverse outcomes, defined as any of the following: in-hospital death, discharge to new residential aged care (not the patient’s usual place of residence), or discharge against medical advice (DAMA). These were chosen to reflect functional decline, transition to dependence, or patient dissatisfaction.

#### Secondary outcomes

##### Measures of hospital resource utilisation

Prolonged hospital LOS, recurrent admissions and need for ICU admission were examined as surrogate measures of healthcare resource utilisation.

Prolonged hospitalisation was defined as hospital LOS above the 90^th^ percentile. Recurrent admissions above 90^th^ percentile were also deemed to need higher healthcare resource usage.

##### Compliance with Hospital Policy

Policies and procedures in our hospital recommend documentation of goals-of-care (GOC), including any treatment limitations, for all patients aged aged 65 years and older, or in situations where clinical judgement suggests that a resuscitation plan is required. GOC documentation is also recommended when patient or Medical Treatment Decision Makers request resuscitation plan discussion, following Medical Emergency Team (MET or Code Blue) activation, for patients of any age with chronic, progressive and life-limiting conditions, patients approaching end of life, patients with multiple co-morbidities and/or at risk of conditions such as stroke or heart failure or cognitive impairment.

To assess adherence, scanned medical records were reviewed for GOC documentation in patients aged ≥65 years and those with high comorbidity burden (calculated from the above comorbidity indices).

### Statistical Analysis

ROC curves were used to evaluate the discriminatory performance of the three comorbidity indices for predicting in-hospital mortality and patient-important adverse outcomes. For each index, the area under the ROC curve (AUC), sensitivity, specificity, precision (positive predictive value), accuracy, and F1 score were calculated. The optimal threshold for each score was determined using the Youden Index.

Hospital LOS, which was not normally distributed, was compared between groups using the Mann–Whitney U test. Descriptive statistics are reported as means (standard deviations) and medians (interquartile ranges) as appropriate.

### Ethics Approval

This study was reviewed and approved by the Mercy Health Human Research Ethics Committee as a negligible risk or quality improvement activity (Study ref: 2023-056). All data were de-identified prior to analysis and securely stored on password-protected hospital computers in accordance with institutional privacy and data governance policies.

## RESULTS

### Baseline Cohort

A total of 21,935 unique adult patients accounted for 29,055 hospital separations between 1 January and 31 December 2022, after excluding patients transferred to other hospitals. The mean age was 45.3 years (SD 18.5), with a median of 38 years (IQR 31–58). The population was predominantly female (17,456/21,935; 79.6%). A summary of cohort characteristics is presented in Supplementary Table 1. Age and sex distributions are shown in Figure 1.

**Figure 1.**
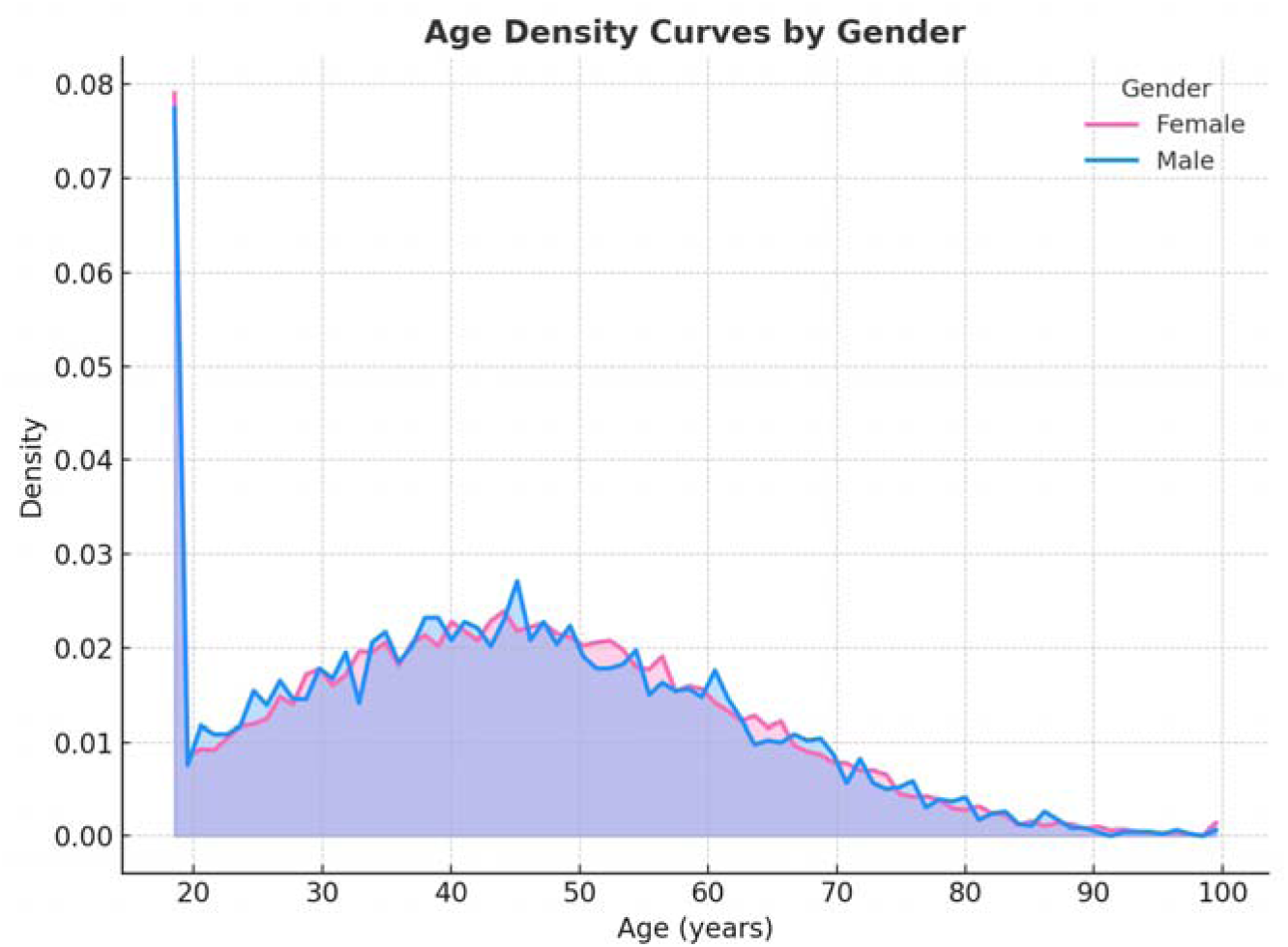
Demographics: gender distribution and age density curve

### Mortality and Patient-important Adverse Outcomes

There were 356 in-hospital deaths, representing a mortality rate of 1.6%. Mortality rates increased with age and differed by gender. When discharges to new residential aged care (n = 74) and discharge against medical advice (DAMA) (n = 176) were included, the composite rate of patient-important adverse outcomes was substantially higher. These outcomes also varied across age groups and between genders. These patterns are illustrated in Figure 2, which displays both in-hospital mortality rates (top panel) and patient-important adverse outcome rates (bottom panel) by age group and gender. Our crude hospital mortality rate was consistent with previously published Australian data [22]. Furthermore, the age-specific mortality profile in our cohort was similar to national mortality patterns reported by the Australian Bureau of Statistics (ABS) for the same period, with mortality rates rising progressively with age and higher among males (Supplementary Figure 3) [23]. This concordance reinforces both the validity and representativeness of our sample.

**Figure 2.**
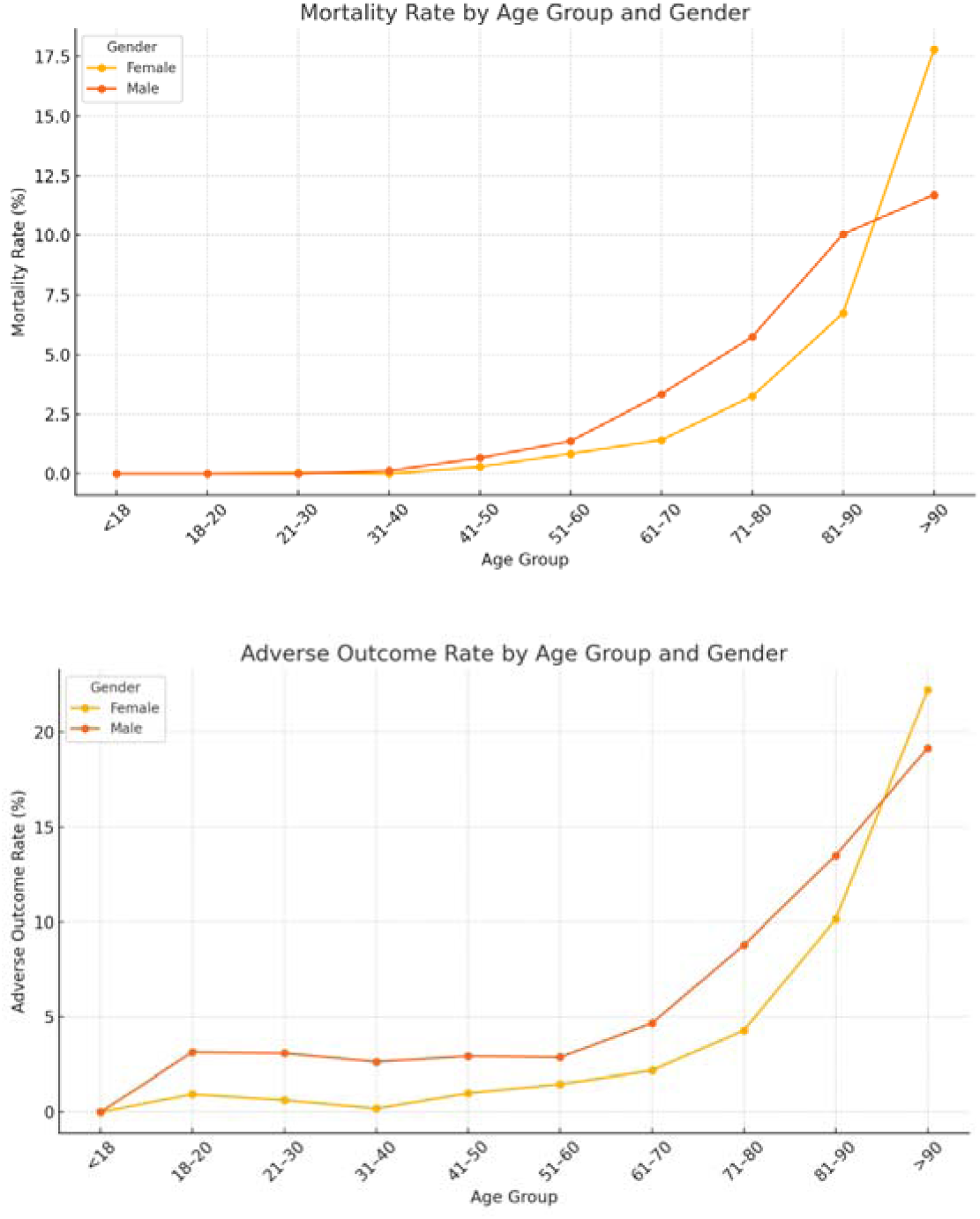
In-hospital mortality and patient-important adverse outcome rates by age group and gender

### Health Resource Utilisation

Patients with hospital LOS ≥ 10days met our definition of prolonged hospitalisation, defined as stays above the 90th percentile. These patients had significantly higher comorbidity burden across all three indices (AHRQ-EI 3 [IQR 0–14] vs 0 [0–0]; VW-EI 5 [0–11] vs 0 [0–3]; age-adjusted CCI 4 [1–6] vs 0 [0–2]; all *p*<0.0001).

Similarly, patients with ≥3 hospitalisations during the study period met the definition of high recurrent admissions (above the 90th percentile). These patients also demonstrated higher comorbidity burden compared with those with ≤2 admissions (AHRQ-EI 0 [0–7] vs 0 [0–0]; VW-EI 0 [0–8] vs 0 [0–0]; age-adjusted CCI 2 [0–5] vs 0 [0–2]; all *p*<0.0001).

The need for ICU admission showed a similar pattern. Patients requiring ICU care had substantially greater comorbidity scores than non-ICU patients (AHRQ-EI 8 [0–14] vs 0 [0–0]; VW-EI 5 [2–12] vs 0 [0–3]; age-adjusted CCI 3 [1–5] vs 0 [0–2]; all *p*<0.0001).

Although ICU admissions accounted for only 2.2% of the cohort (n=491), they consumed 5.2% of all hospital bed-days (4,411/85,406), and ICU patients stayed significantly longer than non-ICU patients (median 5 days [IQR 2–9] vs 1 day [1–3]; *p*<0.0001).

Taken together, these findings demonstrate that higher comorbidity burden is consistently associated with disproportionate use of hospital resources, reflected in prolonged hospital LOS, recurrent admissions and need for ICU care.

### Comorbidity Burden and Predictive Performance

Higher comorbidity burden, as measured by all three indices, was significantly associated with higher rates of adverse outcomes. Discrimination analysis showed that the age-adjusted CCI demonstrated the strongest predictive performance, with an AUC of 0.831, optimal threshold of 4, and recall of 0.702. The VW-EI showed moderate performance (AUC 0.772), followed by the AHRQ-EI (AUC 0.751, threshold 5).

Performance metrics including sensitivity, specificity, precision, and F1 score for all indices are summarised in Table 2. Overall, the aged-adjusted CCI outperformed both Elixhauser-derived scores in discriminating patients at risk of adverse outcomes. The ROC curves and AUC values are illustrated in Supplementary Figure 2.

**Table 2.** Predictive performance of comorbidity indices for adverse outcomes.

| Metric | AHRQ<br>Elixhauser | van Walraven<br>Elixhauser | Aged-adjusted<br>Charlson<br>Comorbidity Index |
| --- | --- | --- | --- |
| AUC | 0.751 | 0.772 | 0.831 |
| Threshold<br>(Youden) | 5 | 5 | 4.0 |
| Accuracy | 0.855 | 0.850 | 0.843 |
| Precision (PPV) | 0.090 | 0.090 | 0.094 |
| Recall (Sensitivity) | 0.601 | 0.622 | 0.702 |
| Specificity | 0.861 | 0.855 | 0.846 |
| F1 Score | 0.157 | 0.157 | 0.166 |

### Goals-of-care (GOC) Documentation

Documentation of GOC discussions was suboptimal even for patients with predictable risk of adverse outcomes. Among patients aged ≥65 years, 49% (n = 1,151) had no recorded GOC documentation. Similarly, 36.5% (n = 3,854) of patients with high comorbidity burden (defined as aged-adjusted CCI ≥4, AHRQ-EI ≥5, or VW-EI ≥5) also lacked documented discussions. The overall GOC distribution across age is shown in Figure 3, while the distribution within the high comorbidity group is presented in Figure 4.

**Figure 3.**
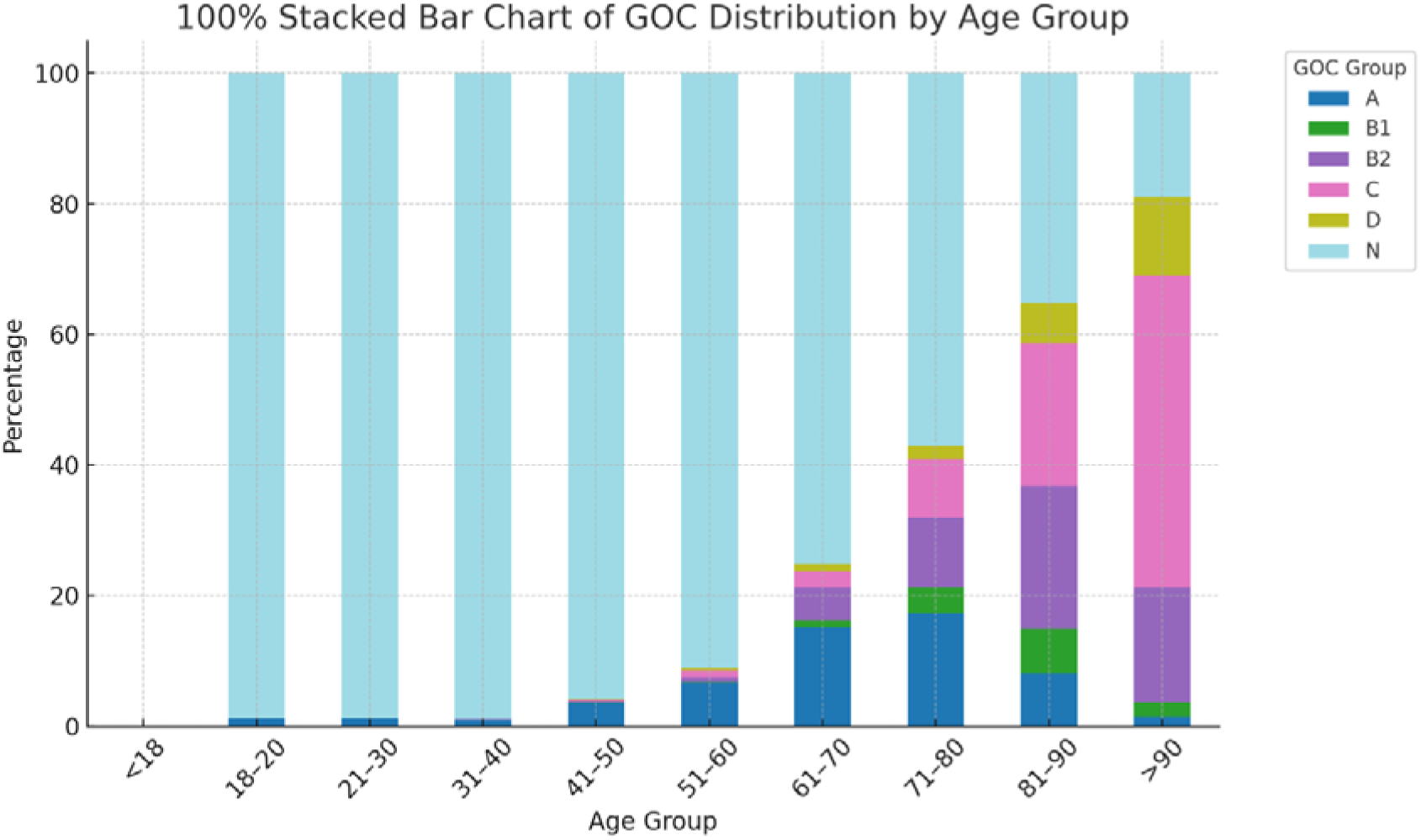
Goals-of-care distribution by age group

**Figure 4.**
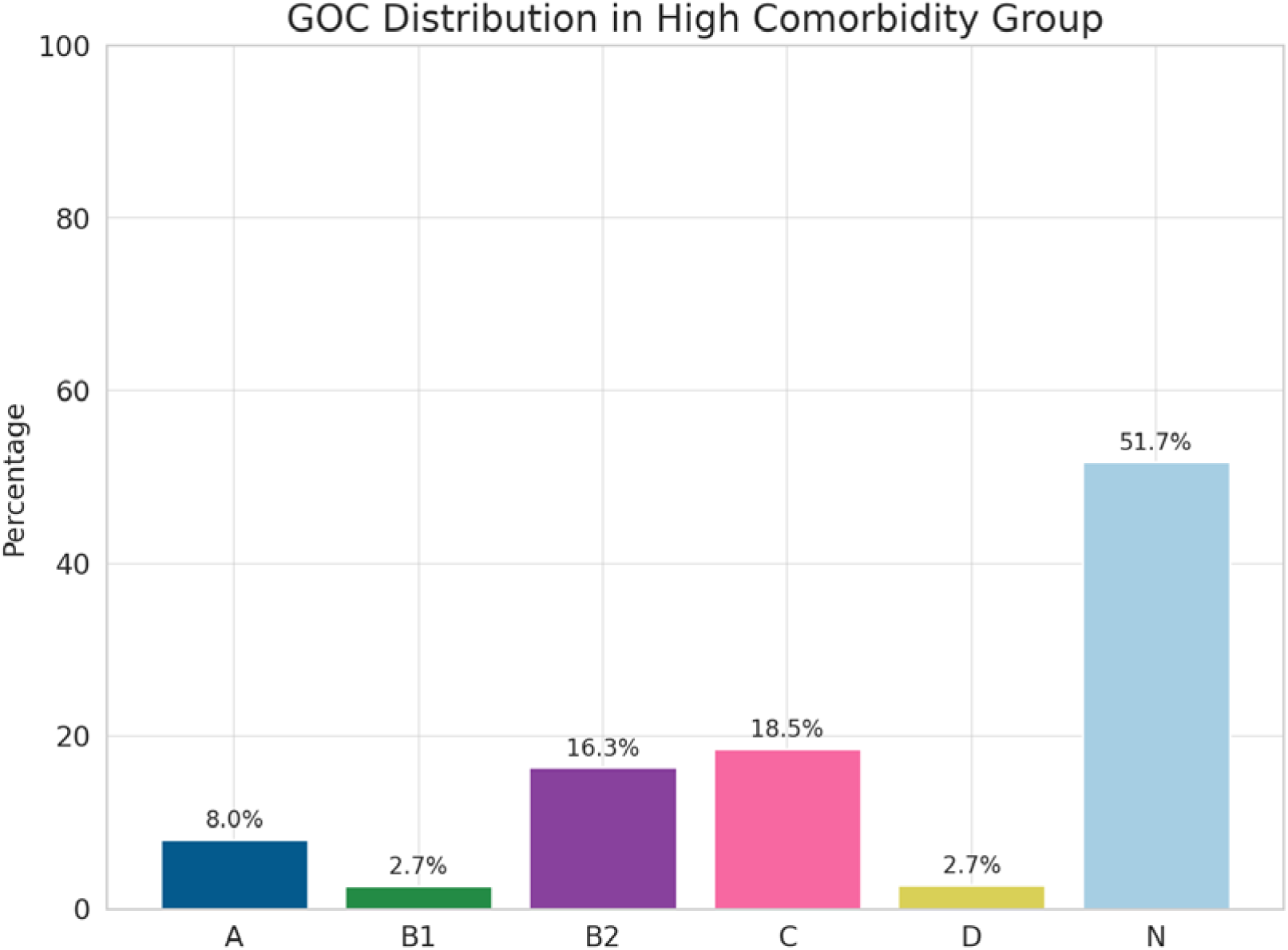
Goals-of-Care (GOC) Distribution in Patients with High Comorbidity Burden

## DISCUSSION

This large, single-centre audit demonstrates that comorbidity burden is a strong predictor of patient-important adverse outcomes and increased utilisation of healthcare resources in the acute care setting. In our cohort, patients with higher comorbidity burden were more likely to experience prolonged hospitalisation, defined as LOS above the 90^th^ percentile (≥10 days), and to have repeated admissions (≥3 hospitalisations during the study period), with both groups showing significantly higher comorbidity scores across all indices (*p*<0.0001). The association extended to intensive care utilisation, where patients requiring ICU admission not only had greater comorbidity burden but also accounted for a disproportionate share of hospital resources, representing just 2.2% of the cohort yet consuming 5.2% of total hospital bed-days, with significantly longer hospital stays compared to non-ICU patients (*p*<0.0001). Taken together, these findings highlight that high comorbidity burden consistently predicts disproportionately high use of healthcare resources, underscoring its potential utility in anticipatory care planning and resource allocation. These findings align with existing literature, including both single-centre studies and systematic reviews, which consistently report associations between increased comorbidity burden and poorer “conventional” healthcare outcomes such as mortality and prolonged LOS across diverse populations and healthcare systems [11–20].

A key strength of this study was the use of a composite endpoint, comprising in-hospital death, new discharge to residential aged care, and DAMA, to better reflect outcomes that matter to patients. While mortality remains a central quality indicator, it alone does not capture broader aspects of health and well-being. Independence, function, and quality of life frequently hold greater value in patients’ preferences [5,6]. Although our composite endpoint is limited by the nature of data collected for an administrative dataset (VAED), these reflect meaningful patient concerns. In addition, our crude hospital mortality closely accords with previously published Australian data [21], supporting the external validity of our findings. The age-related mortality profile observed in our cohort also aligns well with Australian Bureau of Statistics (ABS) national mortality data, suggesting that our sample is representative of the wider Australian population [22].

Among the indices evaluated, the age-adjusted CCI demonstrated the highest discriminative performance (AUC 0.83), followed closely by the VW-EI and AHRQ-EI. These results support a routine role for comorbidity scoring tools to identify high-risk patients in acute care settings and guide early conversations on goals of care aligned with patient values and wishes.

Disappointingly, our findings also reveal persistent gaps in anticipatory care planning. Despite the clear association between age/comorbidity and adverse outcomes, GOC documentation was lacking in over one-third (36.5%) of high-risk patients and nearly half (49.1%) of patients aged ≥65 years. The observed deficit suggests underutilisation of available risk information in routine clinical workflow. Routine comorbidity scoring could support anticipatory care planning, ensure proportionality of treatment, and guide ICU use.

Some limitations should be acknowledged. As with most administrative datasets, reliance on diagnostic coding introduces the risk of misclassification or incomplete comorbidity capture. Patient-reported outcome measures such as health-related quality of life or Functional Independence Measure were unavailable, so proxy endpoints were employed. Being a single-centre, retrospective audit, generalisability may be limited, and causal inferences cannot be drawn. Nonetheless, the findings are consistent with broader literature and suggest meaningful opportunities for improvement in anticipatory care.

Future work should integrate structured, patient-reported outcomes and assess whether embedding comorbidity-driven triggers for anticipatory GOC discussions improves both clinical and experiential outcomes for hospitalised adults. Multicentre validation is also needed to confirm generalisability across diverse healthcare settings.

## CONCLUSION

Comorbidity indices, particularly age-adjusted CCI, are strong predictors of patient-important adverse outcomes and hospital resource utilisation in acute hospital settings. Embedding their use in routine care can support anticipatory goals-of-care discussions, enhance shared decision-making, and help align treatment plans with patient values.

## Authorship

**Dr Hooi Hooi Koay:** First and corresponding author. Responsible for study design, data extraction, statistical analysis, interpretation of findings, and drafting and revising the manuscript.

**Dr Mainak Majumdar**: Senior author and study supervisor. Conceived the study idea, contributed to study design, provided statistical guidance, supervised data interpretation, and critically revised the manuscript for intellectual content.

**Dr Umesh Kadam**: Provided guidance on study design, contributed to ethical review processes, and critically reviewed and revised the manuscript.

**Dr Winnie Theresa** and **Dr Sadeia Shah**: Contributed to data collection, participated in discussions on study design, and reviewed the final manuscript.

## Data Availability

All data produced in the present study are available upon reasonable request to the authors.

## Acknowledgement

The authors would like to thank the Health Information Services team at Werribee Mercy Hospital for their support in providing access to the Victorian Admitted Episodes Dataset. We also acknowledge the contributions of Luke Jennings and Maja Garcia - medical students from The University of Notre Dame Australia, for their valuable assistance with data collection.

## Supplementary Material

**Table 1.** Cohort characteristics.

| Characteristic | Value |
| --- | --- |
| Study period | 1 January 2022 – 31 December 2022 |
| Total separations (admissions) | 30,530 |
| Unique adult patients | 22,720 |
| Mean age (years) | 44.0 (SD 17.9) |
| Median age (years) | 37 (IQR 31–55) |
| Sex, female | 18,071 (80.2%) |
| After deleting external transfer |  |
| Total separations (admissions) | 29,055 |
| Unique adult patients | 21,935 |
| Mean age (years) | 45.3 (SD 18.5) |
| Median age (years) | 38 (31–58) |
| Sex, female | 23,124 (79.59%) |
| Age $\geq 65$ years | 3,870 (17.6%) |
| At least one ICU admission | 491 (2.2%) |
| In-hospital mortality | 365 (1.7%) |
| Discharge to new residential care | 74 (0.3%) |
| Discharge against medical advice (DAMA) | 176 (0.8%) |
| Any patient-important adverse outcome (composite) | 615 (2.8%) |
| High comorbidity burden (CCI $\geq 4$, VW-EI $\geq 5$, AHRQ-EI $\geq 5$) | 4,643 (21.2%) |

**Supplementary Figure 1.**
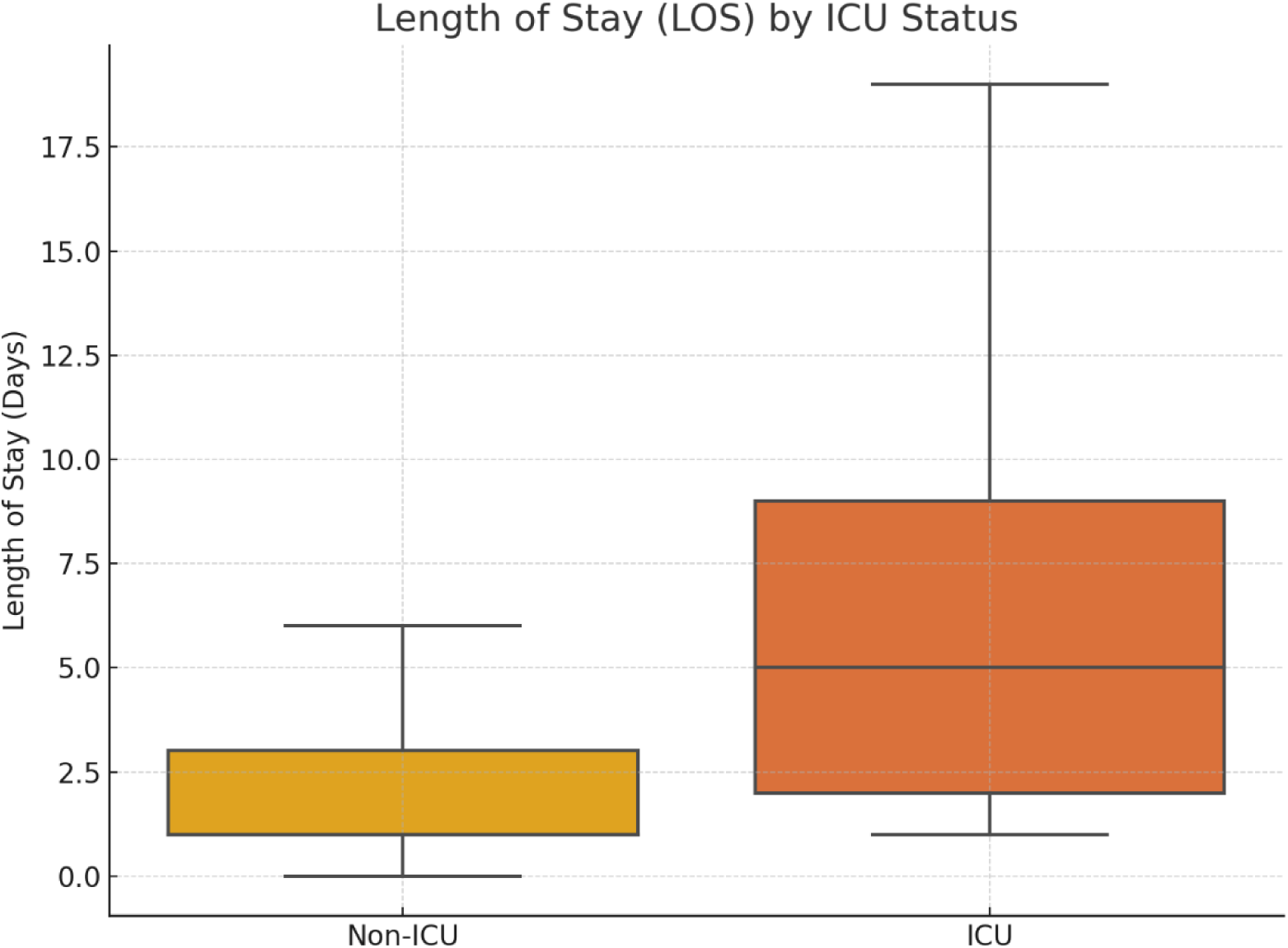
Hospital Length of Stay (LOS) by ICU Admission Status.

**Supplementary Figure 2:**
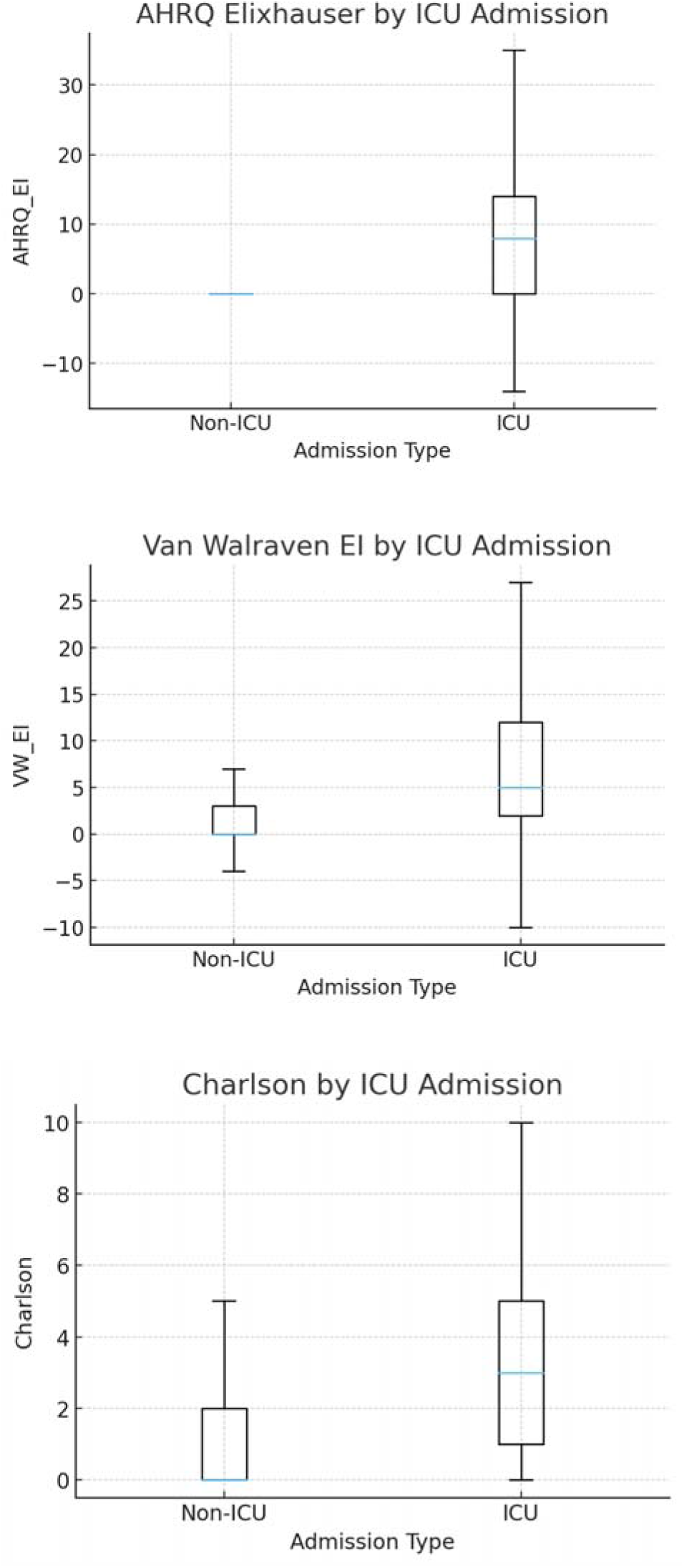
Distribution of comorbidity scores by ICU admission status. Boxplots show (top) AHRQ Elixhauser, (middle) Van Walraven Elixhauser, and (bottom) Charlson comorbidity scores for ICU and non-ICU patients.

**Supplementary Figure 3.**
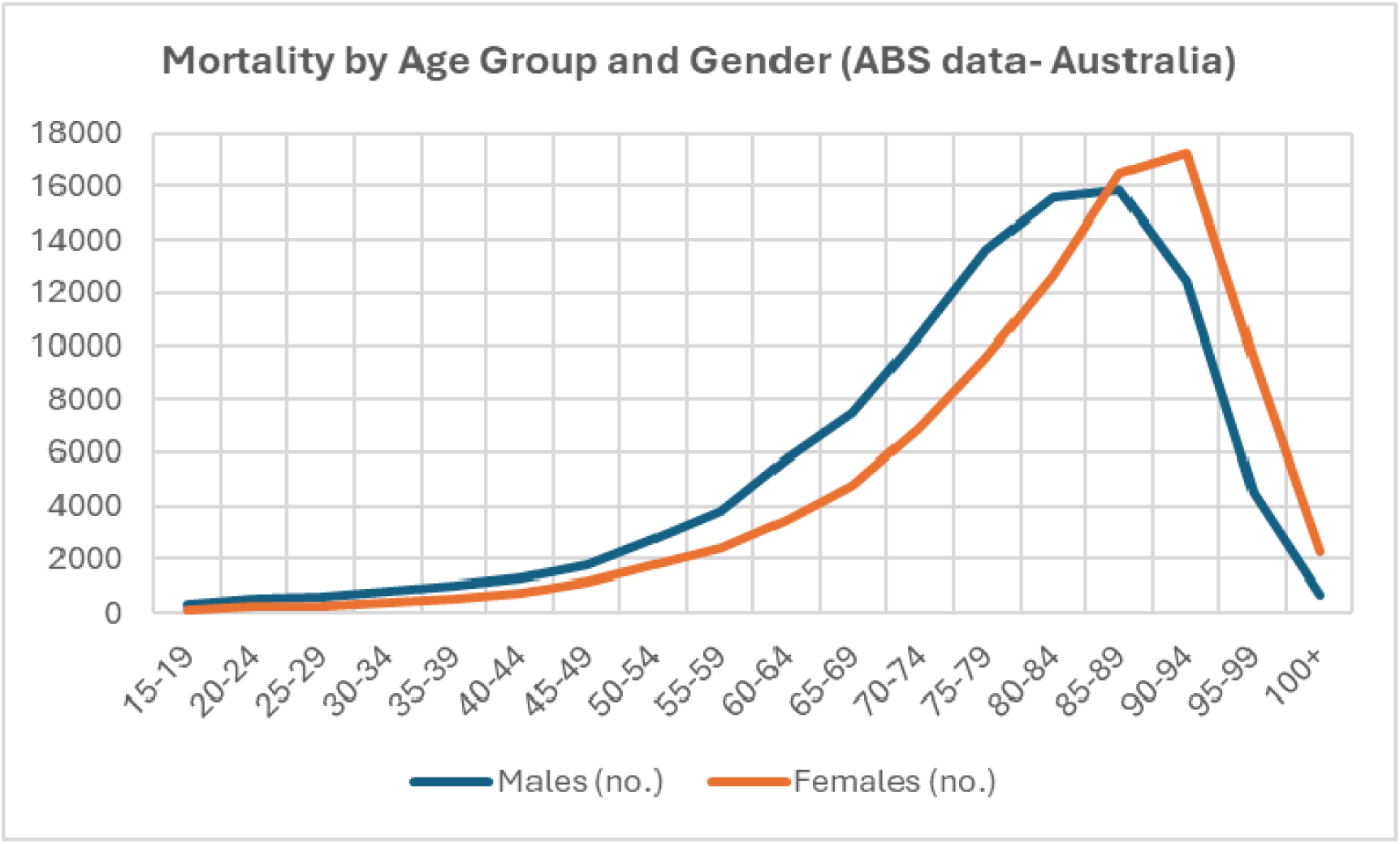

**Supplementary Figure 4.**
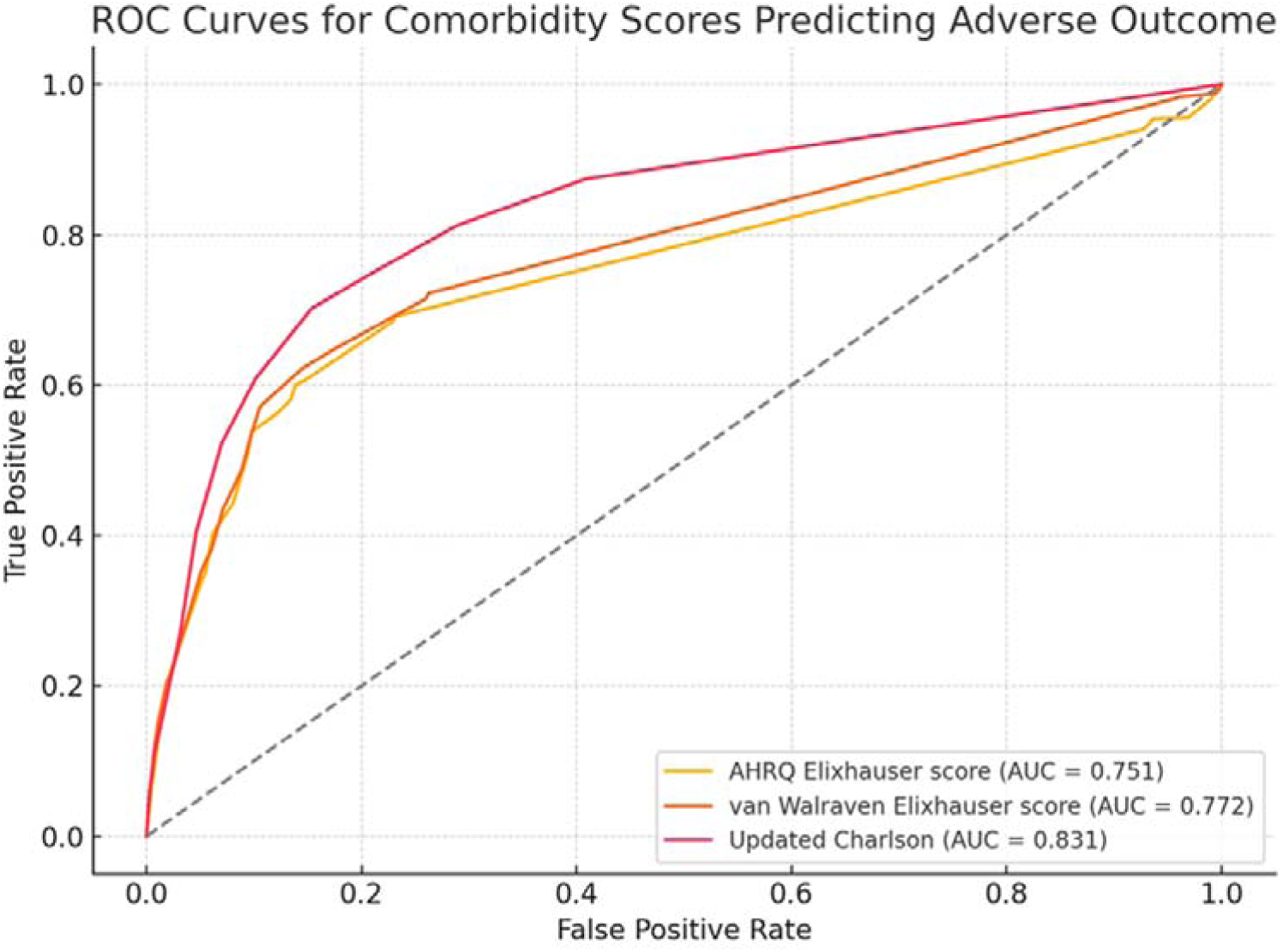
Receiver operating characteristic (ROC) curves for comorbidity indices predicting adverse outcomes

## Notes

### Competing Interest Statement

The authors have declared no competing interest.

### Funding Statement

This study did not receive any funding

### Author Declarations

This study was reviewed and approved by the Mercy Health Human Research Ethics Committee as a negligible risk or quality improvement activity (Study ref: 2023-056).

